# Forecasting hospital demand in metropolitan areas during the current COVID-19 pandemic and estimates of lockdown-induced 2nd waves

**DOI:** 10.1101/2020.07.16.20155721

**Authors:** Marcos A. Capistrán, Antonio Capella, J. Andrés Christen

**Affiliations:** Centro de Investigación en Matemáticas, CIMAT-CONACYT; Instituto de Matemáticas, UNAM, Circuito Exterior, CU, CDMX, Mexico

## Abstract

We present a forecasting model aim to predict hospital occupancy in metropolitan areas during the current COVID-19 pandemic. Our SEIRD type model features asymptomatic and symptomatic infections with detailed hospital dynamics. We model explicitly branching probabilities and non-exponential residence times in each latent and infected compartments. Using both hospital admittance confirmed cases and deaths, we infer the contact rate and the initial conditions of the dynamical system, considering breakpoints to model lockdown interventions and the increase in effective population size due to lockdown relaxation. The latter features let us model lockdown-induced 2nd waves. Our Bayesian approach allows us to produce timely probabilistic forecasts of hospital demand. We have applied the model to analyze more than 70 metropolitan areas and 32 states in Mexico.

## Introduction

The ongoing COVID-19 pandemic has posed a major challenge to public health systems in many countries with the imminent risk of saturated hospitals and patients not receiving proper medical care. Although the scientific community and public health authorities had insight regarding the risks and preparedness measures required at the onset of a zoonotic pandemic, our knowledge of the fatality and spread rates of COVID-19 remains limited [1–4]. In terms of disease handling, two leading issues determining the current situation are the lack of pharmaceutical treatment and our inability to estimate the extent of the asymptomatic infection of COVID-19 [5–7].

Under current circumstances, control measures reduce new infections by limiting the number of contacts through mitigation and suppression [1]. Mitigation includes social distancing, testing, tracing, and isolating infected individuals, while suppression imposes temporary cancellation of non-essential activities. Mitigation and suppression pose a burden on the economy, affecting more individuals living in low-income conditions, challenging the population’s capacity to comply with control measures. As lockdown measures are eased, more people become in contact with the outbreak, and there is a risk of induced 2nd waves that may increase healthcare system pressure.

Broadly speaking, data-driven epidemiological models are built out of the necessity of making forecasts. There are many lessons learned on emergency preparedness and epidemic surveillance from previous pandemic events: AH1N1 influenza [8], MERS [9], SARS [10], Zika [11], Ebola [12], etcetera. However, surveillance data during a pandemic event often suffer from serious deficiencies such as incompleteness and backlogs. Another critical issue is the design of data acquisition, taking into account geographical granularity [13]. Epidemic surveillance of COVID-19 is no different since there is an unknown percentage of asymptomatic infections, and susceptibility is related to economic vulnerability.

Undoubtedly, one key task during the early pandemic response efforts is using epidemiological records and mathematical and statistical modeling to forecast excess hospital care demand with formal quantified uncertainty.

In this paper, we pose a compartmental SEIRD model that considers both asymptomatic and symptomatic infection, including hospital dynamics. We model the residence time in each latent and infected compartments explicitly [14, 15], and we use records of daily confirmed cases and deaths to pose a statistical model that accounts for data overdispersion [16, 17]. Furthermore, we use Bayesian inference to estimate the initial state of the governing equations, the contact rate, and a proxy of the population size to make probabilistic forecasts of the required hospital beds, including the number of intensive care units. The model output has been used by Mexican public health authorities to assist decision making during the COVID-19 pandemic in more than 70 metropolitan areas and the country’s 32 states.

### Contributions and limitations

- We developed a model to produce accurate midterm (several weeks) probabilistic forecasting of COVID-19 hospital pressure, namely hospital beds and respiratory support or mechanical ventilation demands, using confirmed records of cases at hospital admittance and deaths.
- Our model accounts for policy changes in control measures, such as school closures [18] and lockdowns, as breakpoints in the transmission rates.
- Assuming a given fraction of asymptomatic individuals, we infer changes in the transmission rate and the effective population size before and after a given lockdown–relaxation day.
- Inferred changes in effective population size allows us to produce a forecast of lockdown-induced 2nd waves.

Since asymptomatic infection is not fully understood so far [19], the fraction of asymptomatic individuals is yet unknown. Therefore:

- The effective population size is only a proxy, and its absolute value is not meaningful, but only its relative value before and after a relaxation day.
- Without serological studies in the open population – ideally after an outbreak–it is impossible to forecast the population fraction that will be in contact with the virus by the end of the current outbreak.
- At this point, our model does not address next pandemic outbreaks beyond lockdown-induced 2nd waves.
- Finally, although the model does not account explicitly for biases due to behavioral changes [20, 21], population clustering and super spreading events [22], we argue that our approach to lockdowns and relaxation events is a proxy model of these more general events.

### Related work

There are many modeling efforts aimed at forecasting the number of cases, deaths and hospital occupancy during the ongoing COVID-19 pandemic [23–28]. Broadly speaking, models are informed with evolving information about COVID-19 cases, clinical description of the patient residence time in each compartment, fraction of cases per age group, number of deaths, hospital bed occupancy, etc. Columbia University metapopulation SEIR model [23] forecasts are based on assumptions relating an effective contact rate with population density at a metropolitan area and social distancing policies. The COVID Act Now model [24] forecasts the replacement number *R*_*t*_ and the fraction of infections requiring hospitalization using the Bayesian paradigm to fit a SEIR model to cases, hospitalization, death, and recovery counts. The Imperial College response team mathematical model [25] uses an unweighted ensemble of four models to produce forecasts of the number of deaths in the week ahead for each country with active transmission. The IHME model [26] combines a mechanistic model of transmission with curve fitting to forecast the number of infections and deaths. Moghadas *et al*. [27] pose a mechanistic model parametrized with demographic data to project hospital utilization in the United States during the COVID-19 pandemic. The main goal of Moghadas *et al*. is to estimate hospital pressure throughout.

Other COVID-19 models have been used to explore exit strategies [29, 30], the role of recovered individuals as human shields [31], digital contact tracing [32], break points in the contact rate to account for changes in suppression and mitigation policies [18] and lockdown-induced 2nd COVID waves [33] under the assumption that population is temporally geographically isolated.

## Materials and Methods

“Models should not be presented as scientific truth” [34]. Indeed, models are tools intended to serve a specific purpose, evaluate or forecast particular aspects of phenomena and ideally should be developed following the processes of predictive science [35]. Namely, identify the quantities of interest (QoI), verify the computational and mathematical approximation error, including their implication in the estimation of QoI, and calibrate the parameters to adjust the model in light of data to bring it closer to experimental observation. When considering uncertainty, Bayesian inference may be used to calibrate some key model features given data. Finally, a validation process must be used to build confidence in the accuracy of the QoI predictions. Our model is built out of three interrelated components; a law for dynamics, a law for uncertainty, and the choice of parameters.

### Dynamical model

As a proxy of hospital pressure, the quantities of interest in our model are the evolving demand of ICU/respiratory–support beds and no-ICU hospital beds. We developed a full compartmental SEIRD model featuring several compartments to describe hospital dynamics (see Figure 1 and supporting materials, SM) with sub-compartments to model explicitly residence rates as Erlang distributions [14, 15]. The model has two variants, one with age structure and one that assumes age-independent dynamics. Here we describe the latter (see supporting material for some additional comments on the age-dependent model).

**Fig 1.**
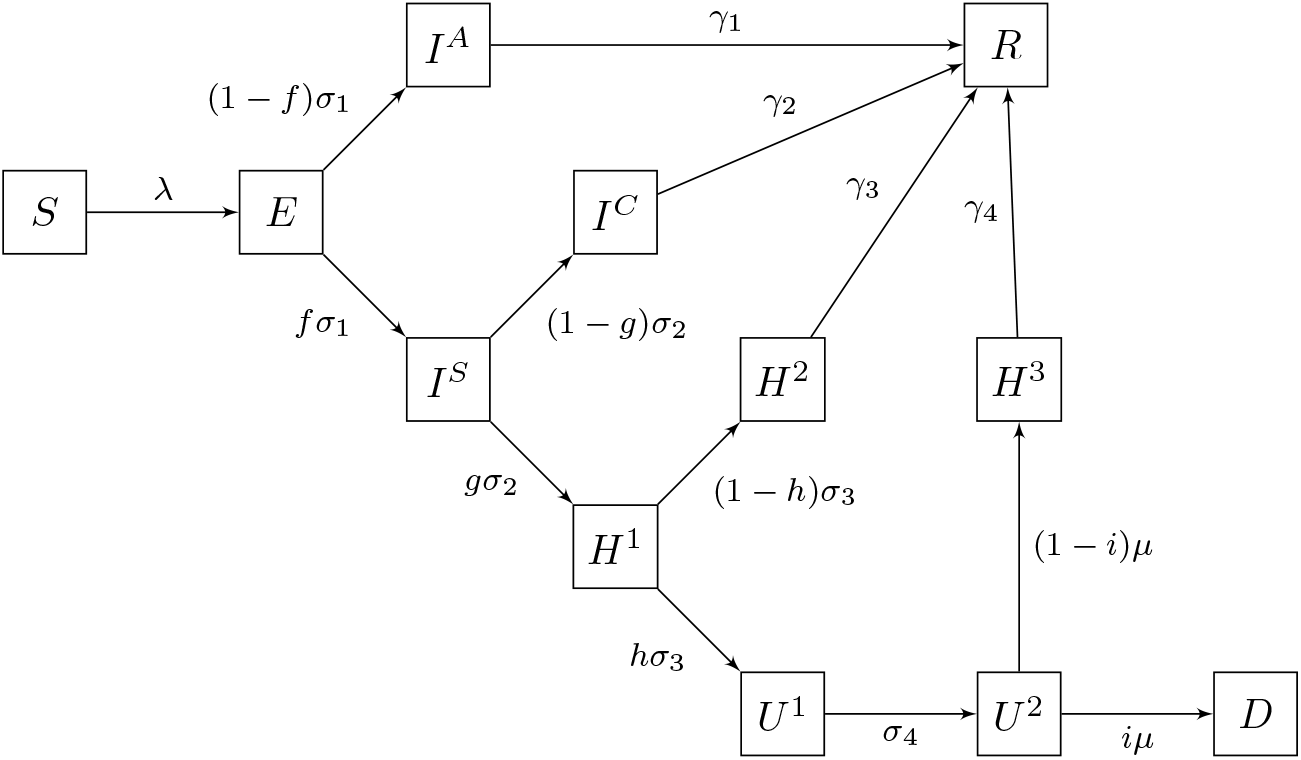
Schematic diagram of the dynamical model. Erlang sub-compartments not shown. For a precise definition of parameters, see supplementary material.

Succinctly our model goes as follows: once the susceptible individuals (*S*) become infected, they remain in the incubation compartment (*E*) for mean time of 1/*σ*_1_ days (i.e. residence rate *σ*_1_). After the incubation period, exposed individuals become infectious and a proportion *f* of them become sufficiently severe symptomatic cases (*I*^*S*^) to approach a hospital, while the remaining cases become mild–symptomatic to asymptomatic (*I*^*A*^). The asymptomatic/mild–symptomatic cases remain infectious a mean time of 1*/γ*_1_ days and eventually recover. For the symptomatic cases (*I*^*S*^) we assume that after an average time of 1*/σ*_2_ days a proportion *g* of infected individuals will need hospitalization (*H*^1^), while the rest (*I*^*C*^) will receive ambulatory care, recovering after an average convalescent time of 1*/γ*_2_ days in quarantine. The hospitalized patients (*H*^1^) remain an average time of 1*/σ*_3_ days until a fraction *h* will need assisting respiratory measures or ICU care such as mechanical ventilation (*U* ^1^). The remaining fraction 1 − *h* of hospitalized patients (*H*^2^) will recover after 1*/γ*_3_ days in average. Respiratory-assisted/ICU patients (*U* ^1^) remain in that state an average of 1*/σ*_4_ days until a critical day is reached when a proportion *i* of them will die (*D*) and the remaining proportion 1 − *i* will recover (*H*^3^) after an average period of 1*/γ*^4^ days. Similar models have been proposed by [2, 31, 32, 36]. For the infection force (*λ*) we assume that only mild–symptomatic/asymptomatic (*I*^*A*^) and symptomatic (*I*^*S*^) individuals spread the infection, that is

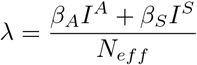

where *β*_*A*_ and *β*_*S*_ are the contact rates of asymptomatic/mild–symptomatic and symptomatic individuals, respectively.

### Parameters and observational model

The model has two kinds of parameters that have to be calibrated or inferred; the ones related to COVID-19 disease and hospitalization dynamics (such as residence times and proportions of individuals that split at each bifurcation of the model) and those associated with the public response to mitigation measures such as the contact rates *β*’s and the effective population size *N*_*eff*_ during the outbreak. Some of these parameters can be estimated from hospital records or found in recent literature or inferred from reported cases and deaths, but some remain mostly unknown. In the latter category, we have *N*_*eff*_ and the fraction 1 − *f* of asymptomatic/mild–symptomatic infections.

Reported values of the proportion of asymptomatic/ mild–symptomatic infections cases 1 − *f* range from 10% to 75%, and even 95% in children population [6, 7, 37]. The number *N*_*eff*_ is lower than the full population of a metropolitan area and depends on different aspects. Still, it is likely to be a consequence of unequal observance of social distancing public policies among the population that, in turn, yield some clustering effects. As lockdown measures are relaxed, more people become in contact with the outbreak, and *N*_*eff*_ may change. This change is a proxy of the fact that the connectivity between clusters increases, and new paths become open to the virus to colonize the full population. Notice that in our model the total number of patients that will visit a hospital is given roughly (bounded) by the product *N*_*eff*_ × *f* and the total number of patients admitted to the hospital is given by *N*_*eff*_ ×*f* ×*g*, where *g* is the portion of infected persons that need hospitalization.

Since our QoI are concerned with hospital pressure, we choose from the available data two sources of information for the observational model: The registered confirmed COVID-19 patients at hospitals, with or without hospitalization, and B deceased patients. Even under an outbreak, these data are reasonably consistent and systematic information on the inflow A and outflow B, hat “hedge” the hospital dynamics. We have evidence (see SM) that given our choice of observation model, the inference of our QoI only depends on the product *N*_*eff*_ ×*f* × *g*, and not on the value of their factors. The fraction *g* is easy to estimate from hospital records (see SM) of admissions and ambulatory patients. Thus we are only required to postulate a value for the product *N*_*eff*_ × *f*.

### Lockdowns, relaxation and lockdown-induced 2nd waves

The model features lockdown intervention and lockdown relaxation as discontinuities in the transmission rate and the effective population size *N*_*eff*_. To model lockdown interventions, a breakpoint is established at which *β* = *β*_1_ before and *β* = *β*_2_ after the intervention day. This creates a non-linear time-dependent *β*(*t*) [18, 38]. In the same fashion, further intervention days may be included by adding more change points and *β* parameters. These additional parameters are then included in the inference.

Assuming that effective population size *N*_*eff*_ and transmission rates are fixed, SEIRD type models converge to the attractor *E* = 0, *I* = 0, i.e. the system models an epidemic that dies out after one single acme. Even for sensible non–constant transmission rates, these kinds of models are only able to produce single epidemic outbreaks. Therefore, in order to be able to estimate secondary outbreak waves after lockdown relaxation measures, one necessarily needs to estimate a different *N*_*eff*_ before and after relaxation days. An increase of *N*_*eff*_ is aimed as a proxy to the rise in the average connectivity between population clusters after relaxation days. Our approach here is as follows: we include the new parameter(s) *ω*_*i*_ *∈* (0, 1) and set *N*_*eff*_ = *ω*_*i*_*N*, where *N* is the total population of the metropolitan area or region under study. Next, we postulate a value of *f* fixed and estimate *ω*_1_ and *ω*_2_ before and after the relaxation day, respectively. To add flexibility to our method and model possible changes in population behavior, we also force a shift from *β*_1_ to *β*_2_ before and after a relaxation day. With this procedure, given *f* we are able to estimate *N*_*eff*_ in terms of the *ω*’s. Nevertheless, due to the confounding effect of the product *N*_*eff*_ × *f* = *N* × *ω* × *f*, we are still unable to estimate the real *N*_*eff*_ ‘s until the real value of *f* is known. Relaxation days may include both lockdown relaxation day or mayor changes in the public response to mitigation measures (see SM for a methodology on how to choose relaxation days).

### Setting lockdown and relaxation days

As explained before, we model interventions and relaxation days as discontinuities in *β* and (*β, ω*), respectively. We consider a lockdown day on 22 March 2020, where a country-wide lockdown started. In Mexico City, we include a second intervention day to model a further local intervention in early April. The methodology to set the relaxation days is as follows: we computed *R*_*t*_ following [39] and look for days where local minimums occur. We choose the relaxation days judiciously, keeping them at least three weeks apart to have enough data to perform the inference. A rise in *R*_*t*_ can be a consequence of an increase in contact rates, effective population size, or both. In either case, our model captures these changes, that serve as a model for the connectivity changes in the outbreak cluster structure.

### Observational model and data

To make our inferences, we use both confirm cases and deceased counts. In some regions, sub reporting of COVID-19 related deaths may become relevant, especially in places hit by a severe outbreak [40]. Nonetheless, deaths are a more reliable data source to estimate a COVID-19 outbreak, especially in the forecast of hospital demand. The problem here is that the number of confirmed cases depends heavily on local practices, particularly with the intensity of testing, adding a level of complication if testing intensity has varied due to ambiguous policies. Regarding data from Mexico, patients are tested when arriving at hospitals with probable COVID-19 symptoms and limited testing is done elsewhere; accordingly, most confirmed COVID-19 cases are limited to A as described above. Therefore, for our inferences, we use both confirm cases A and deceased counts B, as explained in the previous section. Regarding data availability for our observational model, we use the patient’s reported onset of symptoms date. Due to administrative reporting delays, we discard the last 11 days of reporting and add four days as the time stamp for hospital admittance A. We use the registered deceased date as the timestamp for B.

We consider daily deaths counts *d*_*i*_ and its theoretical expectation that is estimated in terms of the dynamical model as *μ*_*D*_(*t*_*i*_) = *D*(*t*_*i*_) − *D*(*t*_*i*−1_) for the metropolitan area or region being analyzed. Analogously, we consider daily cases *c*_*i*_ and its corresponding *μ*_*c*_(*t*_*i*_) given by the daily flux entering the *H*^1^ compartment, which may be calculated as in [17], namely

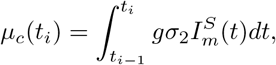

where 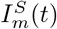 is the last state variable in the *I*^*S*^ Erlang series. We calculate the above integral using a simple trapezoidal rule with 10 points (1/10 day).

### Bayesian Inference

In order to carry out a likelihood-based analysis, we assume that epidemic data has more variation than implied by a standard Poisson process, as is the case in other ecological studies. Following [16], we postulate that the number of both the confirmed cases and deaths follows a negative binomial distribution *NB*. Denoting the mean and variance as *μ* and *σ*^2^, and requiring that *σ*^2^ = *ωμ* + *θμ*^2^ > *μ* we enforce overdispersion for suitable chosen parameters *ω* and *θ*. For data *y*_*i*_, namely *c*_*i*_ and *d*_*i*_, we reparametrize the negative binomial distribution and let *y*_*i*_ ∼ *NB*(*pμ*(*t*_*i*_), *ω, θ*), with fixed values for the overdispersion parameters *ω, θ* and an additional reporting probability *p* (see SM for further details).

We assume conditional independence in the data, and therefore from the NB model, we obtain a likelihood. Our parameters are the contact rate parameter *β*’s, the *ω*’s and crucially we also infer the initial conditions *E*(0), *I*^*A*^(0), *I*^*S*^(0). Letting *S*(0) = *N* − (*E*(0) + *I*^*A*^(0) + *I*^*S*^(0)) and setting the rest of the parameters to zero, we have all initial conditions defined and the model may be solved numerically to obtain *μ*_*D*_ and *μ*_*c*_ to evaluate our likelihood.

Finally, regarding the elicitation of the parameters prior distribution, we use *Gamma* distributions with scale 1 and shape parameter 10 to model the initial conditions *E*(0), *I*^*A*^(0), *I*^*S*^(0) of the community transmission. Following the argumentation of Cori *et al*. [39], we assume that local transmission starts when there are 10 confirmed cases. The rationale for using Gamma distribution priors is that we can specify the distribution by prescribing its first two moments, and the resulting distribution verifies a maximum entropy condition. Namely, we obtain the less informative distribution that has the prescribed mean and (log) variance [41]. The prior for the first transmission rate *β*_0_, is a long tail, log Normal with *σ*^2^ = 1 and scale parameter 1; that is *log*(*β*_0_) ∼ *N* (0, 1). For the subsequent *β*’s, we use autoregressive priors to impose some coherence from one change point to the next with *log*(*β*_*i*_) ∼ *N* (*log*(*β*_*i*−1_), 1). The prior on *ω*_*i*_ is a *Beta*(1 + 1/6, 1 + 1/3) restricted to *ω*_*i*_ > *ω*_*i*−1_. This beta distribution is a fairly flat near uniform density in [0, 1], touches zero in 0 and 1, and is slightly skewed to lower values. We model here the unlikely values *ω*_*i*_ = 0, 1 and that under current social distancing measures we expect smaller rather than larger *N*_*eff*_. Otherwise, the prior for the *ω*_*i*_’s is rather diffuse and non-informative.

To sample from the posterior we resort to MCMC using the “t-walk” generic sampler [42]. The MCMC runs semi-automatic, with consistent performances in most data sets. For any state variable *V*, the MCMC allows us to sample from the posterior predictive distribution for *V* (*t*_*i*_). By plotting some of its quantiles sequentially, we may produce predictions with quantified probabilistic uncertainty, as seen in Figure 3.

**Fig 2.**
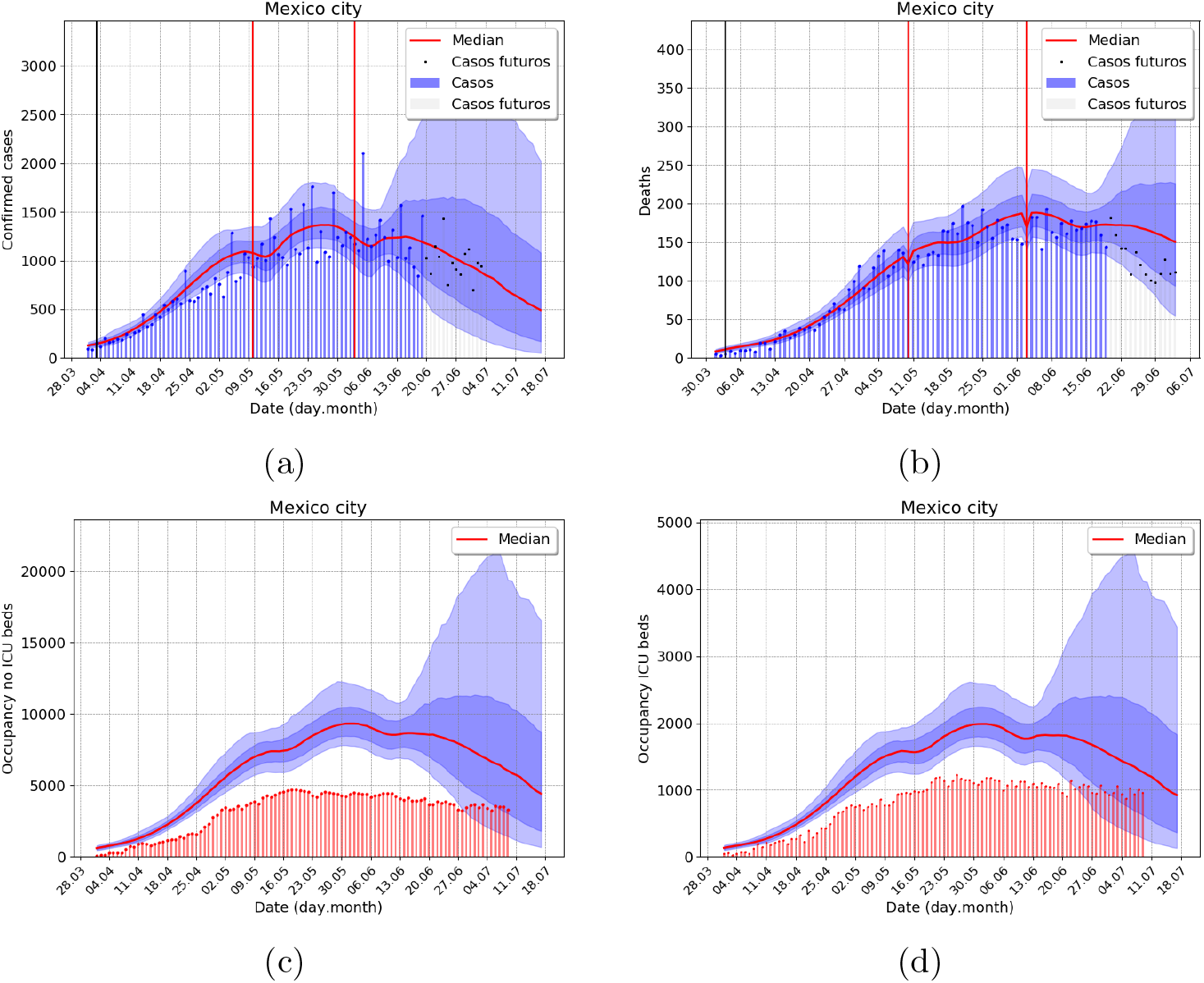
Outbreak analysis for Mexico city metropolitan area, using data from 7 July 2020. Gray bars correspond to two weeks of trimmed data. (a) Incidence of confirmed cases, (b) Incidence of deaths (c) No ICU, and (d) ICU demand of hospital beds. Total population 21, 942, 666 inhabitants

**Fig 3.**
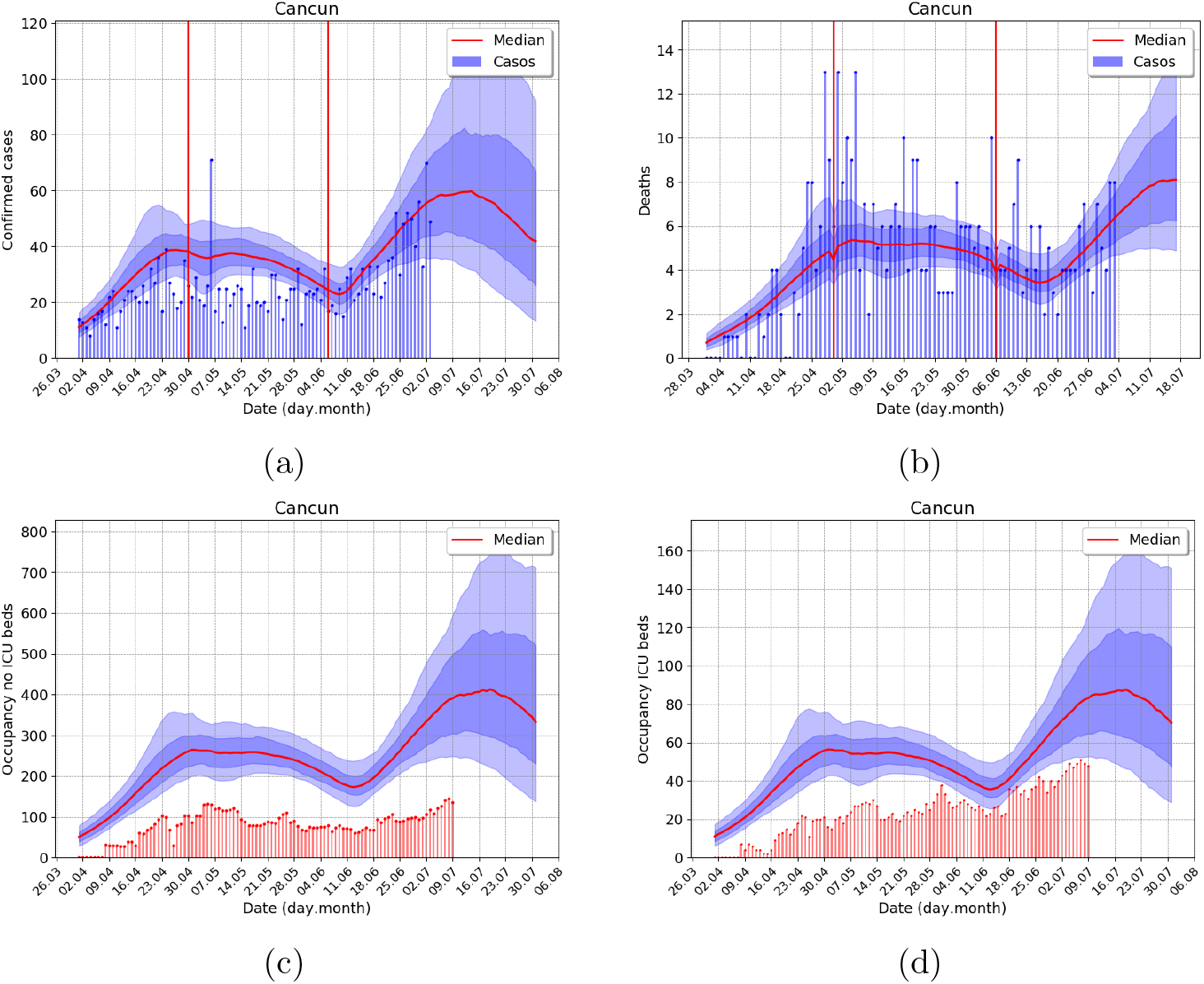
Outbreak analysis for Cancun metropolitan area, using data from 7 July 2020. (a) Incidence of confirmed cases, (b) Incidence of deaths (c) No ICU, and (d) ICU demand of hospital beds. Total population 891, 843 inhabitants.

As in the case of climate forecasting, due to the stochastic nature of a pandemic outbreak point-wise estimates such as the maximum a posteriori estimate (MAP) does not provide good descriptions of the outbreak evolution. No single trajectory of the SEIRD model provides a good description of the outbreak evolution, nor give accurate forecasts.

## Results

Local transmission started at different dates in each Mexican metropolitan area given the different communicability with Mexico City and with the rest of the world. On the other hand, a country-wide general lockdown started on May 22nd until June 1st where each state started differently local control measures. Figure 3 (a) shows the model forecast, with quantified uncertainty, of the daily records of COVID-19 confirmed cases in Mexico City. Gray bars correspond to two weeks of trimmed data to assess the model performance.

Figure 3 (b) depicts records and forecasts incidence of deaths. In Figure 3 (c) and (d) we compare the model forecasts with hospital bed and ICU occupancy obtained from a secondary official source of epidemiological surveillance depicted as red bars. Notice that the forecast begins after June 20th, and an uncertainty cone opens to the right the next 3 to 4 weeks. However, the attractor of the dynamical system closes the cone for longer times, and the predictive power of our forecast decreases.

The estimate of total hospital bed occupancy corresponds to the daily integral of *H*^1^ and *H*^2^ in the model and the ICU occupancy corresponds to the *U* ^1^ daily integral. We consistently overestimated the total number of hospital beds and ICU units and shifted to earlier times. We calibrate residence times from reports on daily demand of hospital beds and intensive care unit records from *Instituto Mexicano del Seguro Social* or Mexican Social Security Institute (IMSS) at the early stages of the outbreak. Given the virus’s emerging nature and evidence of an excess of hospital pressure in places like Spain, Italy, and New York. We deliberately assumed value parameters to forecast higher demands in both types of hospital occupancy.

In Figure 3 (a) and (b), black and red vertical lines represent lockdown and relaxation days, respectively. Our model forecasts three different bump-shape regions where the effective population size increases.

### Mexico city, metropolitan area

### Cancun metropolitan area

We present the case of Cancun’s metropolitan area since it is a medium-sized city with considerable international connectivity was among the first ones with an outbreak in Mexico. The forecast shows a clear first wave, with a long decreasing tail and a lockdown-induced 2nd wave after a lockdown easing and reopening of touristic activities.

In Figure 4 we show the three posterior distributions for *ω*. In (a), we show the Mexico City case, and in (b), we show the Cancun metropolitan area case. In the supplementary material, we show the outbreak analysis for some other cities to illustrate different aspects of our forecasting model’s performance. Besides this paper’s examples, we apply our model to 70 metropolitan areas and the 32 states in Mexico (“ama” model; https://coronavirus.conacyt.mx/proyectos/ama.html, in Spanish).

**Fig 4.**
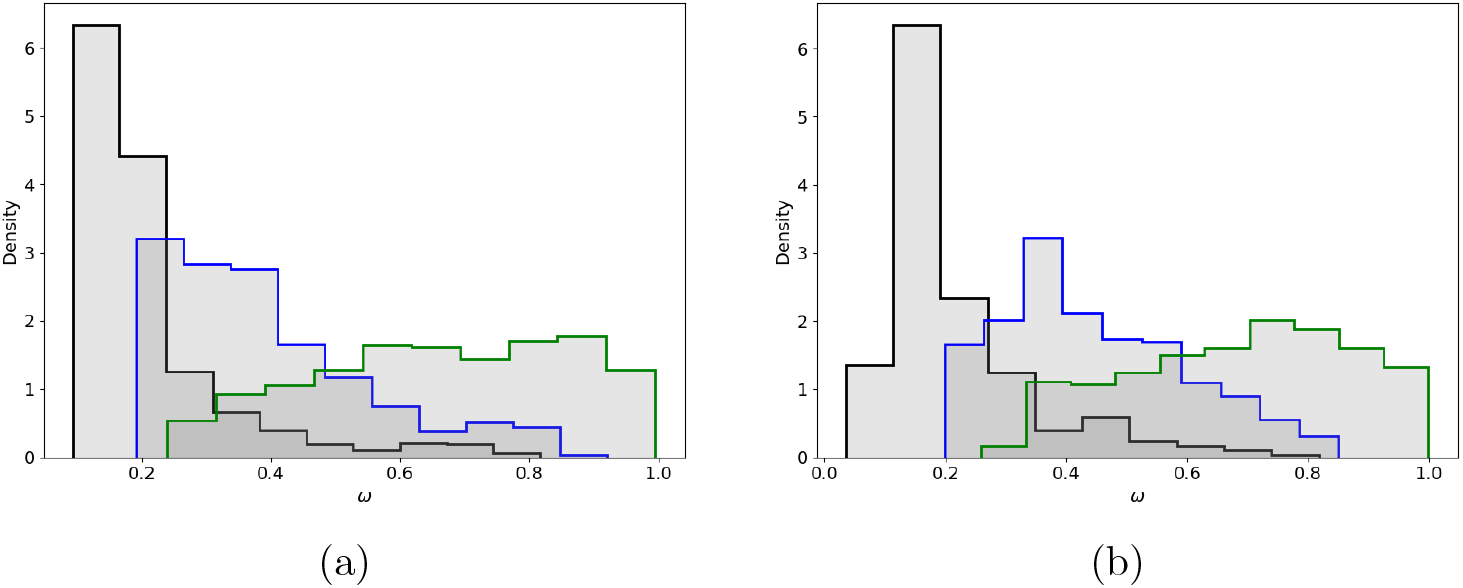
Posterior distribution for *ω*’s (*N*_*eff*_ = *Nω*_*i*_, *f* = 0.4) (a) Mexico city (b) Cancun.

## Discussion

We present a compartmental SEIRD model to make probabilistic forecasts of hospital pressure during COVID-19 outbreaks in metropolitan areas. In this work, we also consider lockdowns and lockdown-relaxations as two different kinds of interventions. We model the former as a change in transmission rates while the latter also allows for changes in the effective population size. These changes in effective population size and transmission rates are used as a proxy of behavioral changes, changes in the connectivity between population clusters and super spreading events.

We in-line monitor exogenous changes in SEIRD parameters to inform the model of interventions and relaxation days, in the same spirit of our inferences in the transmission rate and initial conditions. Our analysis showed that we account for the outbreak evolution in many metropolitan areas by setting one lockdown on 22 May 2020, the beginning of country-wide lockdown, and two more relaxation days. One around 10 May, mother’s day, a popular celebration in Mexican culture where families gather together. The other relaxation day is set around four to eight days after 1 June, the announced date for the country-wide lockdown relaxation measures. In some cases, we also impose somewhat different relaxation days to account for local changes, such as the opening of tourist activity. Of note, all relaxation days where set exclusive by our *R*_*t*_ based methodology. The interpretation, as above, came afterward.

In a prior version of this ama model –before reaching the first acme– we did not infer the effective population size, namely *ω*. Instead, by modeling several countries that already had passed the acme, we postulated *ω* = 1 and *f* = 0.05. This yields some controversy that even reached the news media [43]. With the current model we can show that the product *ω* × *f*, that is the quantity that affects the estimates, takes maximum a posteriori values between 0.04 and 0.12 in most of our studied cases, see Figure 6. This fact explains the reason why our forecast weeks ahead on the acme have been rather accurate. It is also important to notice that this is evidence of a confounding effect between *ω*,, and the transmission rates *β* on the early stages of the outbreak. Many forecasting models that fail to recognize this fact will consequently fail on their acme estimates.

**Fig 5.**
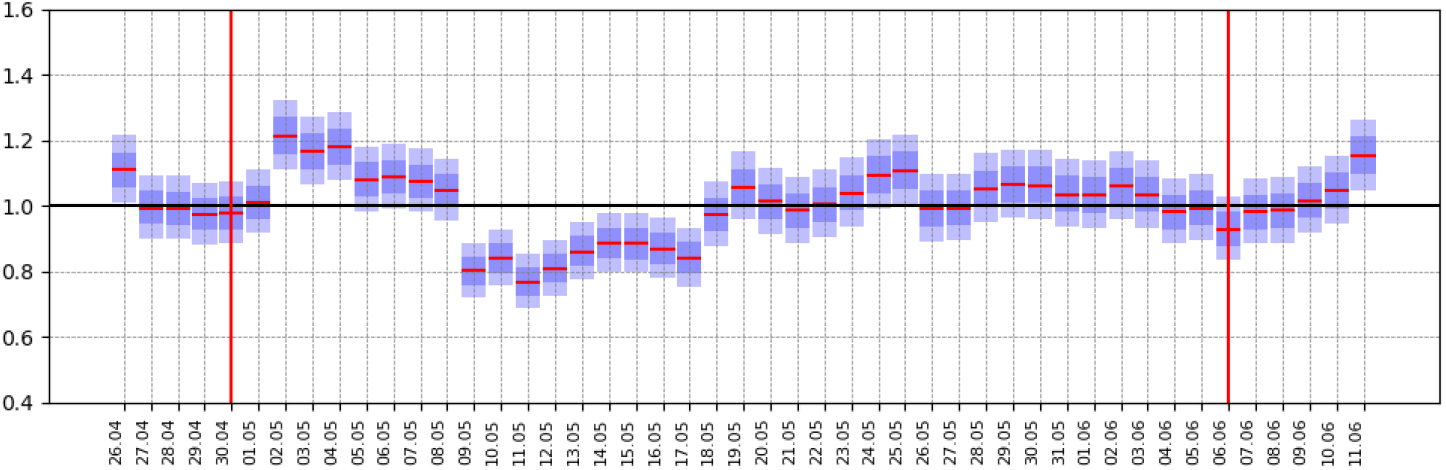
Daily *R*_*t*_’s calculated as in [39] for Cancun metropolitan area. Two relaxation days, marked with red vertical lines, were judiciously included at local minimums, allowing for a minimum gap of 3 weeks.

**Fig 6.**
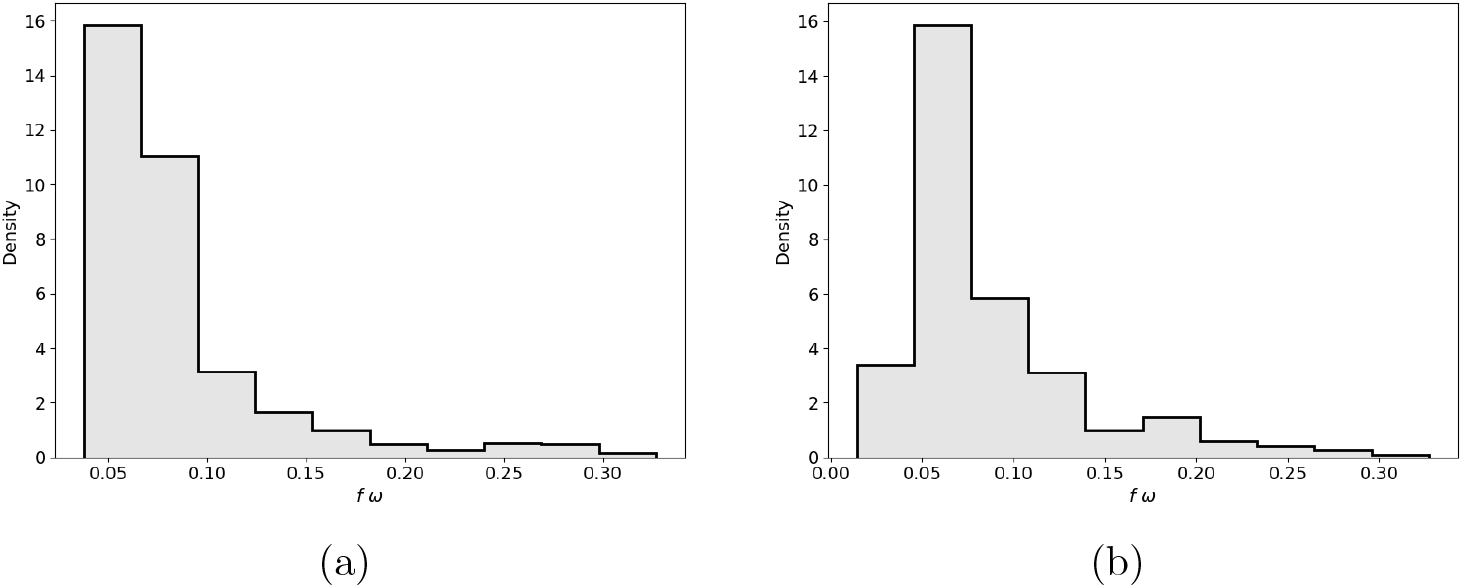
Posterior distribution of *f* × *ω*_0_ for (a) Mexico city and (b) Cancun metropolitan areas, as explained in Section. Note how, using the proxy value of *f* = 0.4, the proportion *f* × *ω*_0_ is estimated close to 0.05. In a previous version of this model we postulated *ω* = 1 and *f* = 0.05.

We note that after the acme SEIRD type models converge to the attractor *E* = 0, *I* = 0, and long term estimates tend to overestimate the decline of an outbreak. The underlying assumption that social contact and other conditions remain constant is not reasonable for most societies over long periods (e.g., more than eight weeks). Conservative short to mid-term forecasts most be preferred and change points added when necessary.

The age-independent model has proven to be adequate to produce accurate forecasts for the hospitalization dynamics during current outbreaks. Therefore the added complexity of the age-structure model may not justify its use at this point. However, we continue to work in our age structure model.

Since our QoI are related to the hospital pressure, we choose all parameters conservatively. However, Erlang densities can not correctly approximate residence times of some cases real distributions and more general distributions should be considered. Moreover, as health professionals learn to treat the disease, hospital residence times also change. Both these effects should also be considered to obtain more accurate estimates.

Our observation model is designed to integrate data after the nonlinear term in the flow diagram of the dynamic model (see Figure 1) and the rest of the dynamics is proportional to the hospital occupancy curves, therefore the model forecasts can be used as a proxy of the full outbreak. We follow this idea to predict the date when an outbreak will reach its first peak as well as lockdown induced second waves.

The confounding effect between the population size, namely *ω*, and the fraction of asymptomatic/mild–symptomatic infections 1 − *f* makes it impossible to forecast the population that will be in contact with the virus at the end of an outbreak reliably. Likewise, although it is possible to make a model based analyses of scenarios of lockdown exit strategies, scenario estimation is limited due to the lack of information regarding population viral seroprevalence. Therefore, without serological studies in the open population after a COVID-19 outbreak it is not possible to assess the final outbreak size.

Up to our knowledge, there are very few models that can produce forecast lockdown-induced 2nd waves with quantified uncertainty. Although more elaborate models can be considered, our model is simple and flexible enough to deliver reliable and useful forecasts.

## Data Availability

We shall make all data available upon publication

https://coronavirus.gob.mx/datos/

## Acknowledgments

The authors wish to thank Elena Álvarez-Buylla, Paola Villarreal (CONACYT) and Hugo López-Gatell (Secretaría de Salud) for their support and comments for improving this forecast model and also Instituto Mexicano del Seguro Social (IMSS) for sharing data to adjust the hospitalization dynamics. Special thanks to CIMAT technicians Judith Esquivel-Vázquez and Oscar González-Vázquez (CIMAT-CONACYT), for their help in retrieving data and running our programs at a CIMAT cluster. We also thank the CONACYT COVID-19 response group, for additional comments in the development of our model. The authors are partially founded by CONACYT CB-2016-01-284451 grant. AC was partially supported by UNAM PAPPIT–IN106118 grant.

## Supporting information

### S1 Other examples

#### S1.1 Toluca

**Fig S1.**
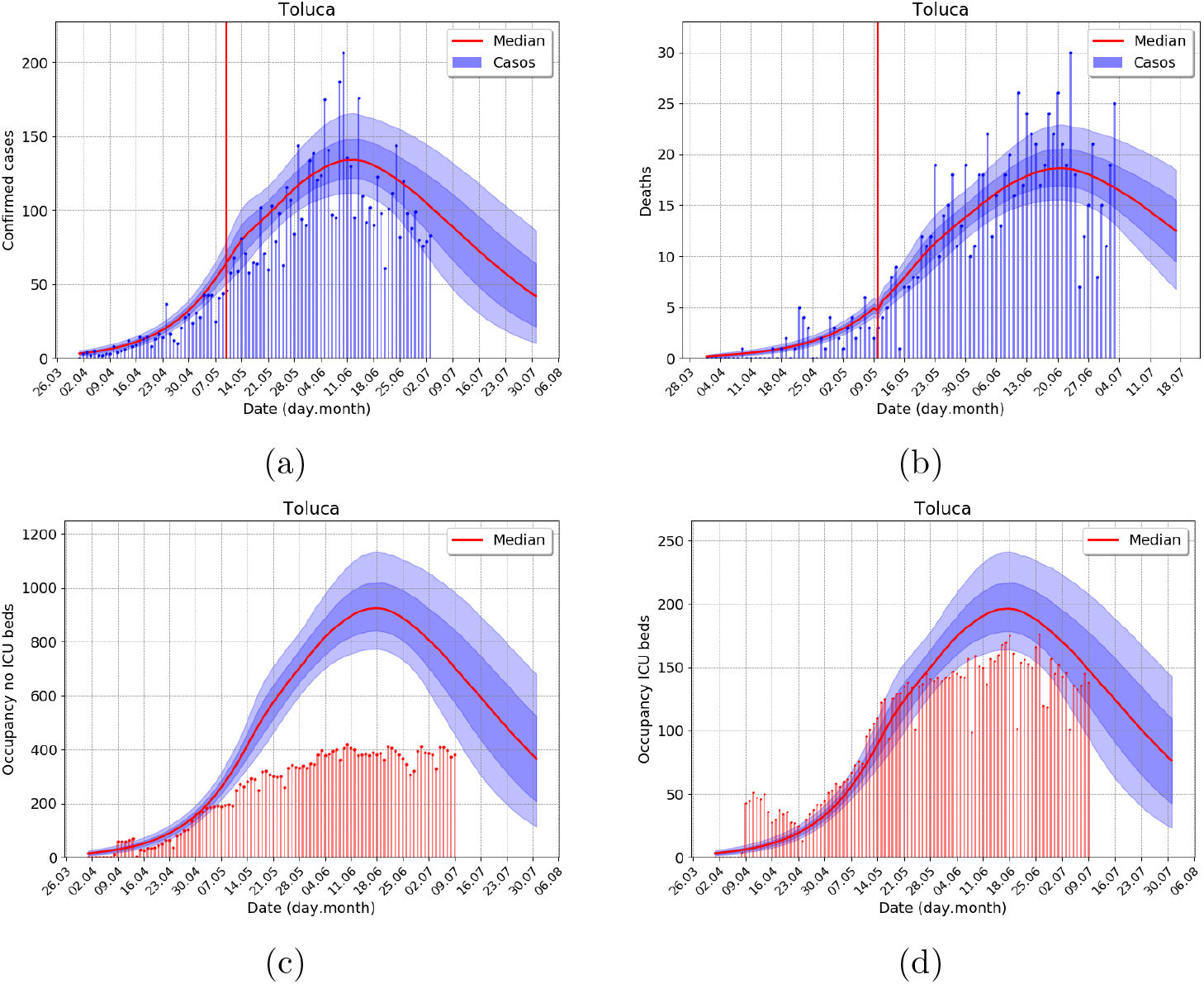
Outbreak analysis for Toluca (the state of Mexico, Mexico central highlands) metropolitan area, using data from 7 July. (a) Incidence of confirmed cases, (b) Incidence of deaths (c) No ICU, and (d) ICU demand of hospital beds. Total population 2, 377, 828 inhabitants. Forecast for the city of Toluca is in course with one relaxation event.

#### S1.2 Merida

**Fig S2.**
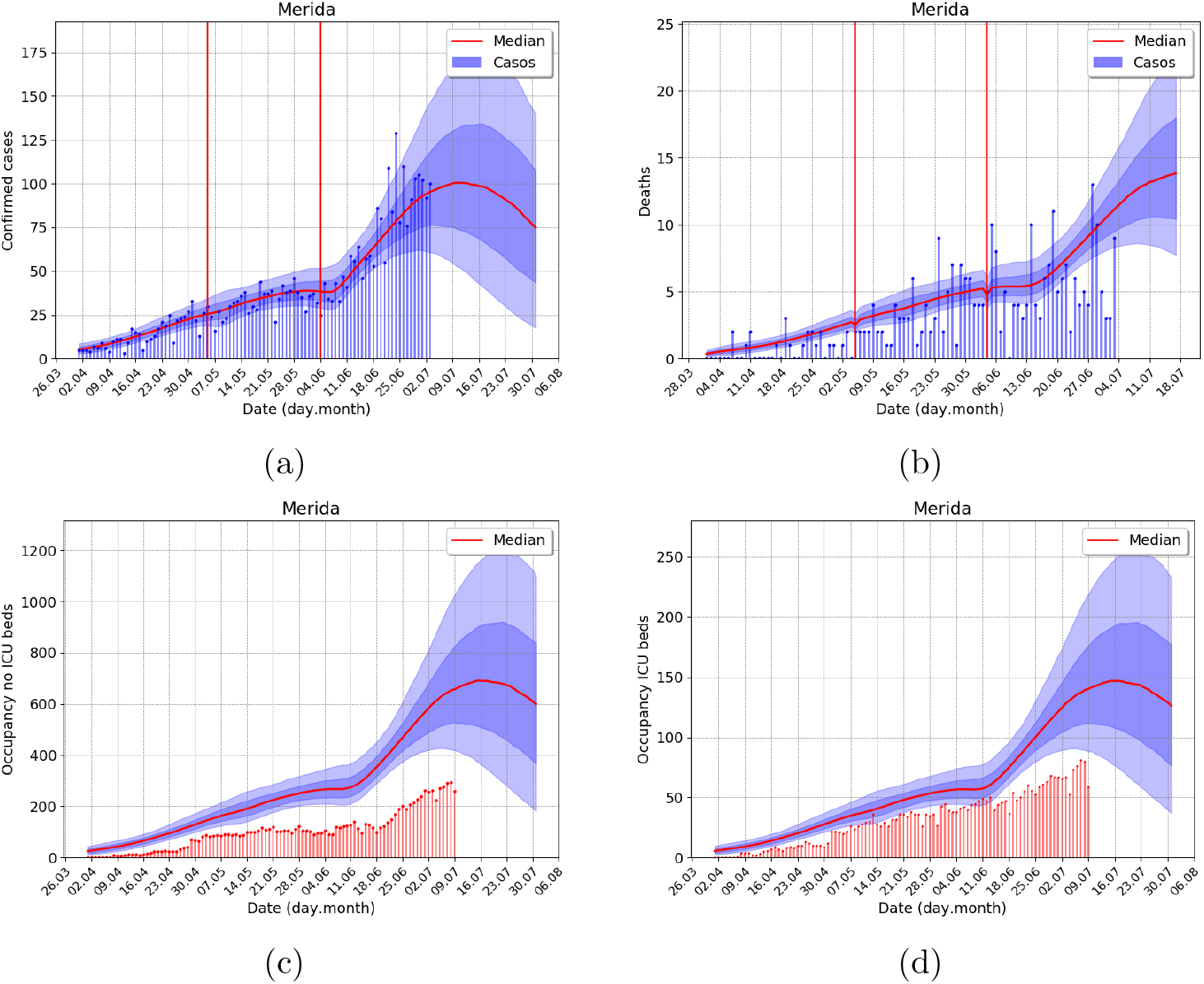
Outbreak analysis for Merida (state of Yucatan, Yucatan peninsula) metropolitan area, using data from 7 July. (a) Incidence of confirmed cases, (b) Incidence of deaths (c) No ICU, and (d) ICU demand of hospital beds. Total population 1, 237, 697 inhabitants. There is an evident ongoing second outbreak in the city of Merida.

#### S1.3 Cuernavaca

**Fig S3.**
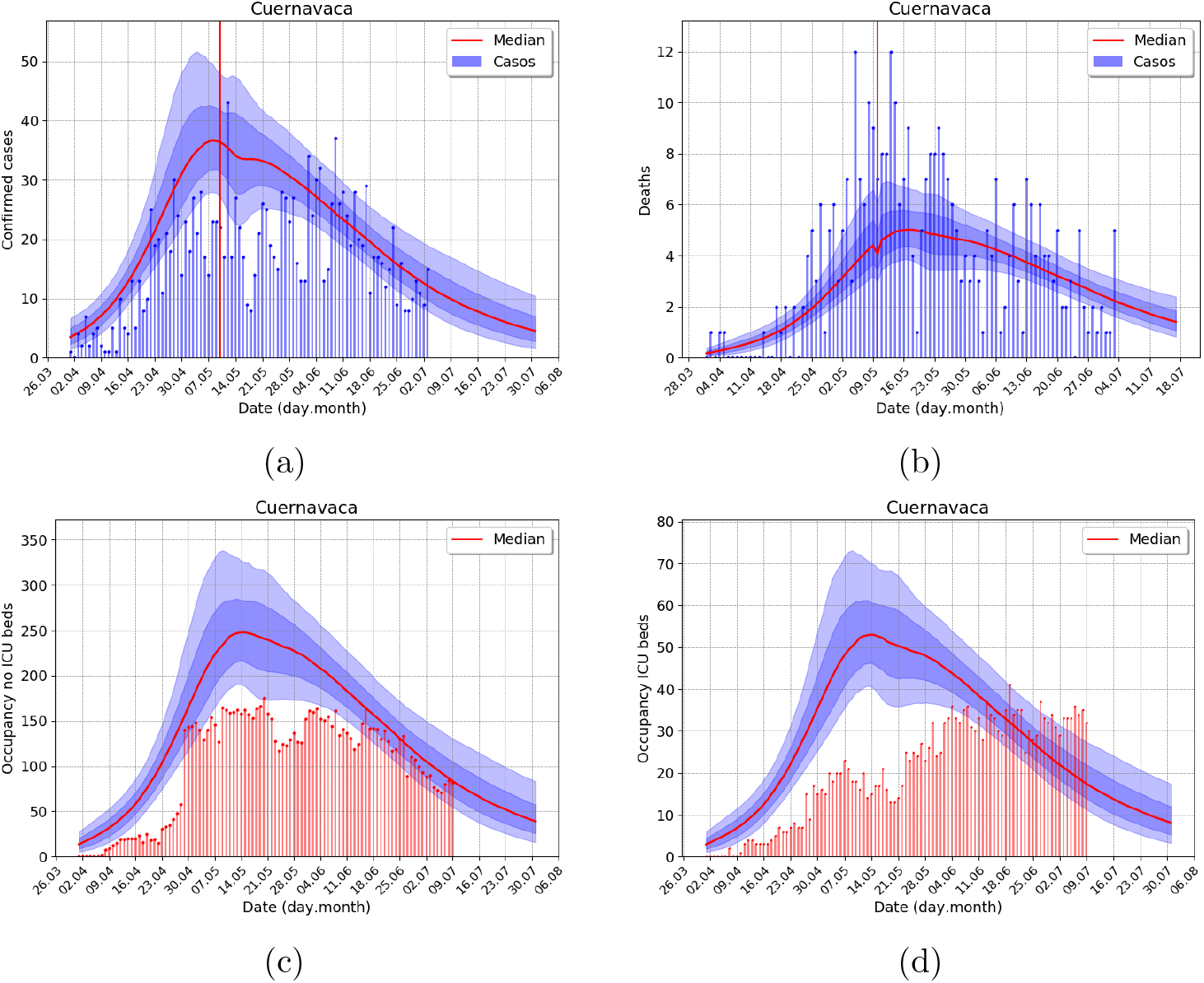
Outbreak analysis for Cuernavaca (the state of Morelos, Mexico central highlands) metropolitan area, using data from 7 July. (a) Incidence of confirmed cases, (b) Incidence of deaths (c) No ICU, and (d) ICU demand of hospital beds. Total population 1, 059, 521 inhabitants. In the city of Cuernavaca, the model captures the slow decline of the outbreak.

#### S1.4 Acapulco

**Fig S4.**
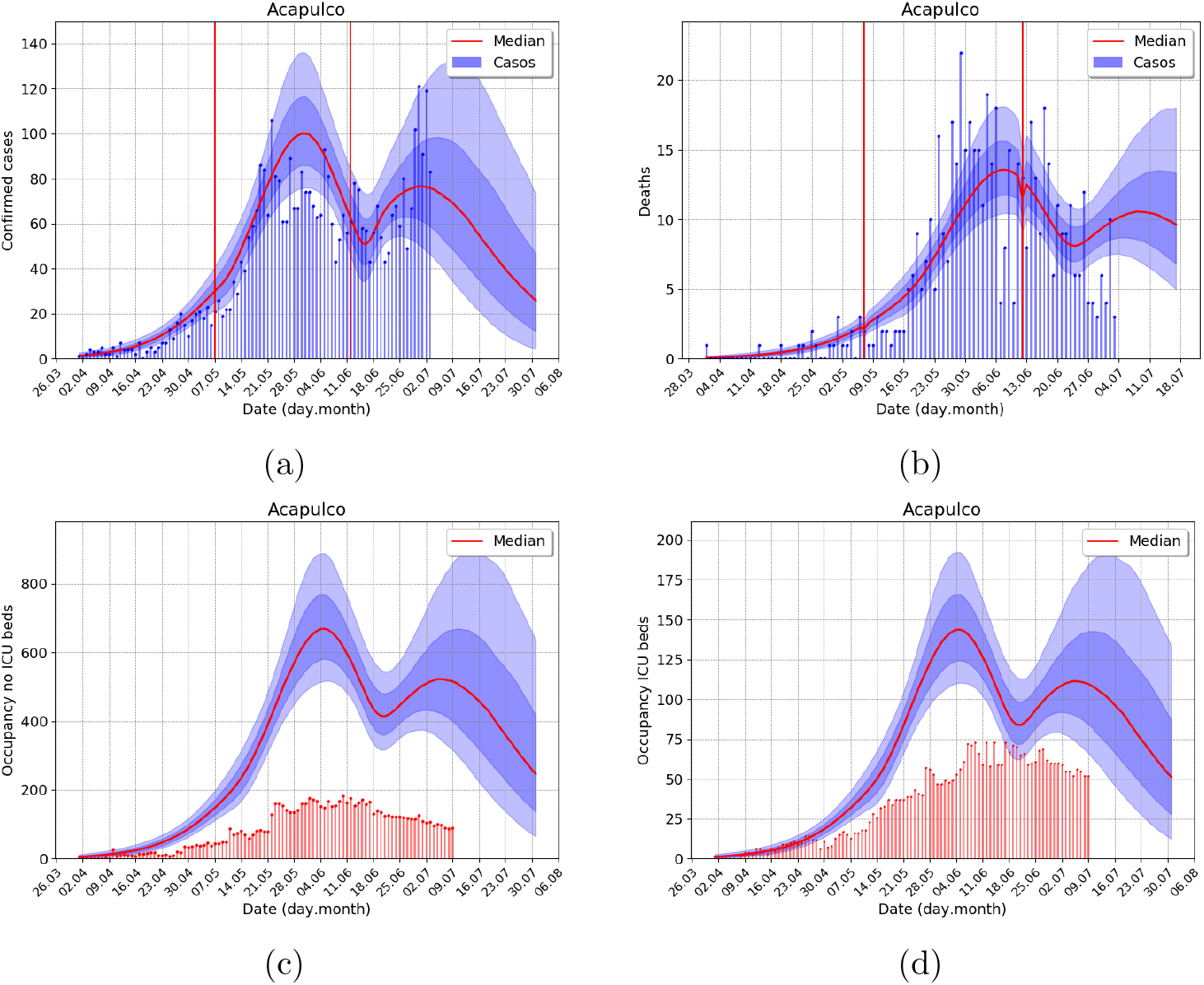
Outbreak analysis for Acapulco (the state of Guerrero, Mexico south pacific shore) metropolitan area, using data from 7 July. (a) Incidence of confirmed cases, (b) Incidence of deaths, (c) No ICU, and (d) ICU demand of hospital beds. Total population 1, 059, 521 inhabitants. The outbreak in the city of Acapulco is an example of a second outbreak of the same size.

#### S1.5 Culiacan

**Fig S5.**
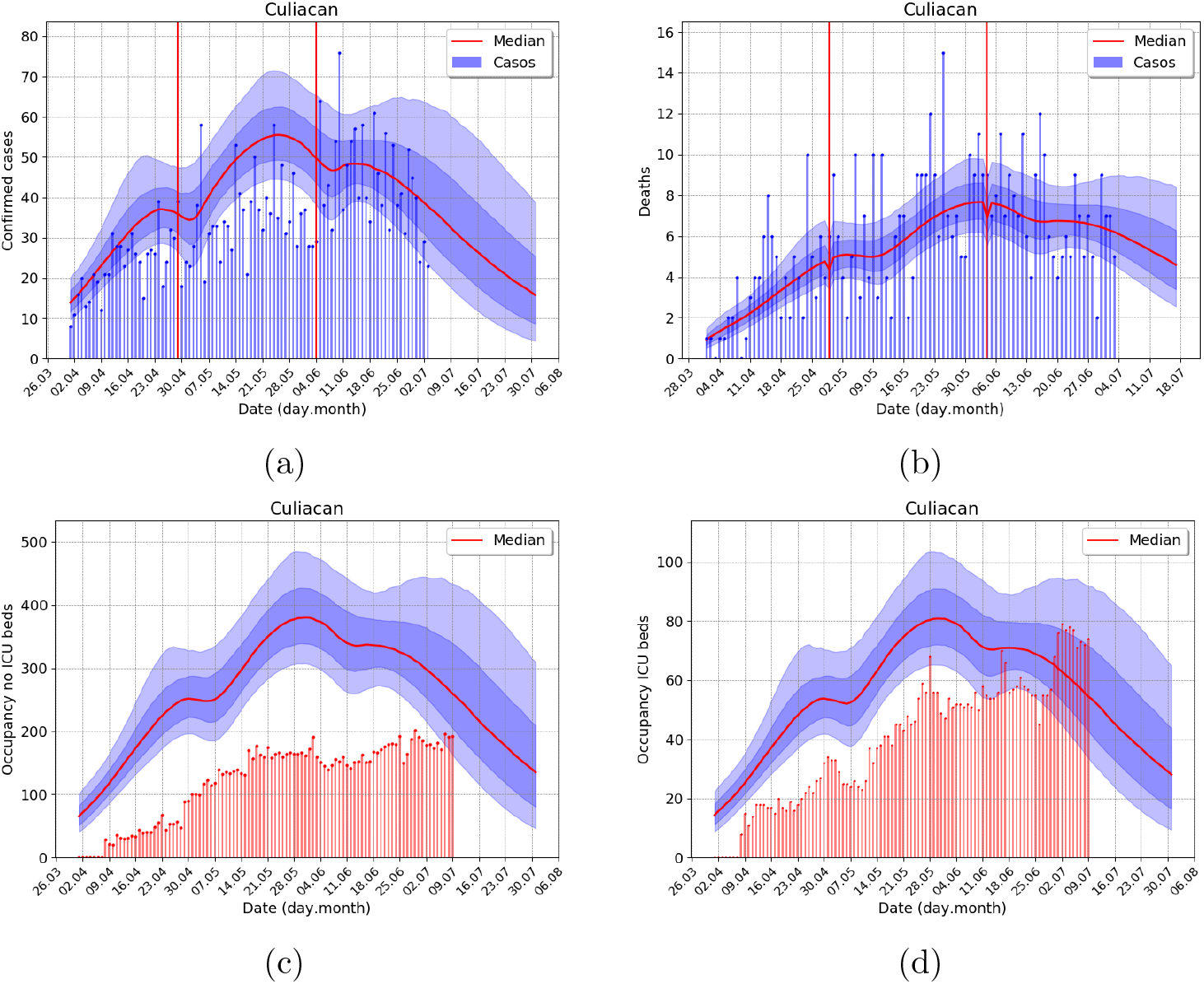
Outbreak analysis for Culiacan (state of Sinaloa, Mexican north pacific shore) metropolitan area, using data from 7 July. (a) Incidence of confirmed cases, (b) Incidence of deaths, (c) No ICU, and (d) ICU demand of hospital beds. Total population 962, 871 inhabitants. Example of secondary outbreak with two lockdown-induced second waves.

### S2 Model

We developed a dynamic transmission compartmental model to simulate the spread of the novel coronavirus SARS-CoV-2. A definition of the state variables is given in Table S1. Additionally, an “Erlang series” is included for most of these state variables to account for non-exponential residence times. The model may be described conceptually with the graph in Figure 1. Without showing the Erlang series for the sub-compartments the system of equations in the model is as follows:

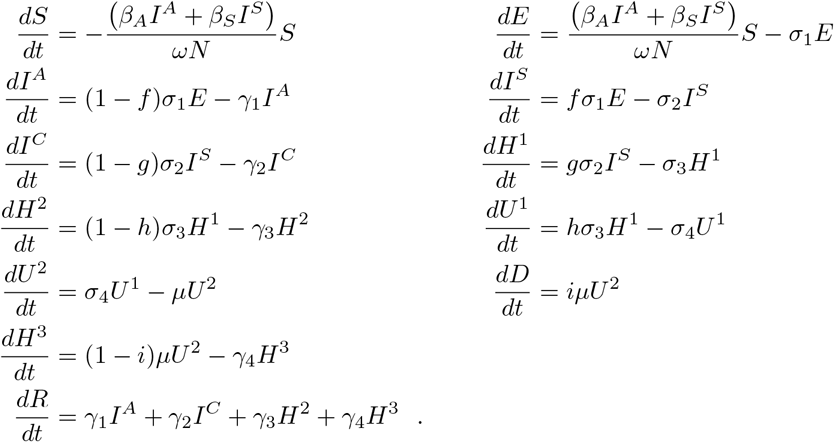

**Table S1.**
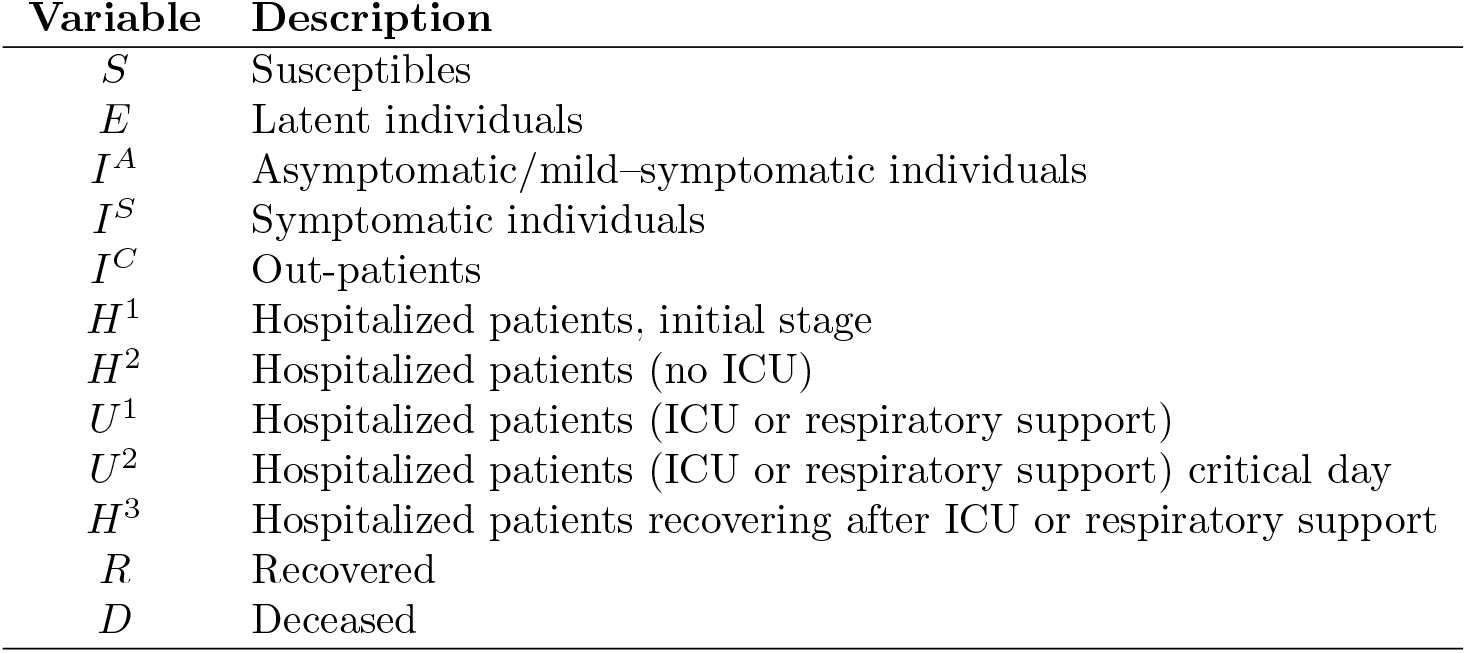
Description of the state variables in the dynamic model

| Variable | Description |
| --- | --- |
| $S$ | Susceptibles |
| $E$ | Latent individuals |
| $I^A$ | Asymptomatic/mild-symptomatic individuals |
| $I^S$ | Symptomatic individuals |
| $I^C$ | Out-patients |
| $H^1$ | Hospitalized patients, initial stage |
| $H^2$ | Hospitalized patients (no ICU) |
| $U^1$ | Hospitalized patients (ICU or respiratory support) |
| $U^2$ | Hospitalized patients (ICU or respiratory support) critical day |
| $H^3$ | Hospitalized patients recovering after ICU or respiratory support |
| $R$ | Recovered |
| $D$ | Deceased |

In Table S2, we give a brief description of all the parameters in the model.

#### S2.1 Infection force and basic reproductive number ℛ_0_

For the infection force (*λ*) we assume that individuals that spread the infection correspond to the mild–symptomatic/asymptomatic (*I*^*A*^) and symptomatic individuals (*I*^*S*^) before they get contact with the health care system or doctor, e.g.

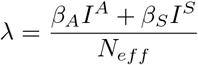

We compute the basic reproductive number *R*_0_ of the epidemic by the next generation matrix method [44] and obtain

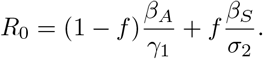

#### S2.2 Values of model parameters

Since our QoI are related to the hospital pressure we choose all parameters conservatively. For each metropolitan area, we assume that *N* corresponds to its full population, as defined by Instituto Nacional de Estadistica y Geografia (INEGI). The values of the transition probabilities are summarized in Table S3.

**Table S2.** Description of model parameters.

| Parameter | Description | Units |
| --- | --- | --- |
| $N$ | Total number individuals in the population | – |
| $\omega(t)$ | Percentage susceptible individuals in the population at time $t$ | – |
| $\beta_A(t)$ | Transmission rate of asymptomatic/mild-symptomatic individuals (asx/mild-sym) at time $t$ | per day |
| $\beta_S(t)$ | Transmission rate of symptomatic individuals at time $t$ | per day |
| $\kappa$ | Relative strength between the transmission the rate of asx/mild-sym and symptomatic | – |
| $f$ | Proportion of infected persons with strong enough symptoms to visit a hospital | – |
| $g$ | Proportion of infected persons that need hospitalization | – |
| $h$ | Proportion of hospitalized patients requiring respiratory support or ICU care | – |
| $i$ | Proportion of Respiratory-assisted or ICU patients deceased | – |
| $1/\sigma_1$ | Average incubation time | day |
| $1/\sigma_2$ | Average time from symptomatic onset to hospital visit | day |
| $1/\sigma_3$ | Average time from hospital admission to respiratory support or ICU care | day |
| $1/\sigma_4$ | Average time with respiratory support or ICU care | day |
| $1/\mu$ | Average length of critical stage of respiratory-support/ICU between death and recovery | day |
| $1/\gamma_1$ | Average time that asymptomatic/mild-symptomatic individuals remain infectious | day |
| $1/\gamma_2$ | Average time of symptomatic individuals that recover without visiting a hospital | day |
| $1/\gamma_3$ | Average time from hospital admission to hospital discharge | day |
| $1/\gamma_4$ | Average time from respiratory-support/ICU care release to hospital discharge | day |

**Table S3.** Transition probabilities at bifurcations in the model

| Parameter | Value | Reference |
| --- | --- | --- |
| $f$ | 0.40 | postulated |
| $g$ | 0.04375 | [1], IMSS |
| $h$ | 0.25 | [1], IMSS |
| $i$ | 0.5 | [1], IMSS |

#### S2.3 Erlang series and sub-compartments

To make the intrinsic generation-interval of the renewal equation in each compartment more realistic we divide each compartment of the model into *m* equal sub-compartments to generate an Erlang–distributed waiting time [14]. The Erlang distributions of each compartment is calibrated by two parameters: the rate *λ*_*E*_ and the shape *m*, a positive integer that corresponds to the number of sub-compartments on the model. In terms of these parameters the mean of the Erlang distribution is *m/λ*_*E*_, this mean correspond to the average times in the dynamic model.

**Table S4.** Average times and Erlang shape parameters for sub-compartments

| Variable | Rates | Average time | Erlang shape $m$ | Reference |
| --- | --- | --- | --- | --- |
| $S$ | $\beta_S$ | Inferred | 1 | — |
| $E$ | $1/\sigma_1$ | 5 days | 4 | [2] |
| $I^A$ | $1/\gamma_1$ | 7 days | 3 | [45] |
| $I^S$ | $1/\sigma_2$ | 4 days | 3 | [46] |
| $I^C$ | $1/\gamma_1$ | 7 days | 3 | [47] |
| $H^1$ | $1/\sigma_3$ | 2 days | 10 | [3] |
| $H^2$ | $1/\gamma_3$ | 10 days | 3 | [3] |
| $U^1$ | $1/\sigma_4$ | 10 days | 3 | IMSS |
| $U^2$ | $1/\mu$ | 1 day | 1 | [47] |
| $H^3$ | $1/\gamma_4$ | 4 days | 5 | [3] |
| $R$ | None | — | — | — |
| $D$ | None | — | — | — |

We use recent publications and information generously shared by the Instituto Mexicano de Seguridad Social (IMSS) to estimate the average time and the shape parameter of the Erlang series in each compartment. In Table S4, we give details of Erlang series lengths, residence times and imputed values.

#### S2.4 Relative strength between the transmission the rate of asymptomatic/mild–symptomatic and symptomatic

In our methodology, we aim to infer the force of the infection *λ*. This parameter is defined in terms of contact rate of asymptomatic/mild–symptomatic individuals *β*_*A*_ and contact rate of symptomatic individuals *β*_*S*_. Due to the functional dependence of *λ* in these parameters, there is a lack of identifiability between *β*_*A*_ and *β*_*S*_ that can not be resolved without further assumptions. We assume that the relative strength between the transmission rate of asymptomatic/mild–symptomatic and symptomatic is modeled as a fixed ratio *κ*. We model the value of *κ* directly as the ratio of the viral load of symptomatic and asymptomatic/mild–symptomatic patients [48, 49] and fixed it to *κ* = 2. Hence, the force of infection becomes *λ* = *β*^*S*^(*I*^*S*^ + *κI*^*A*^)*/N*_*eff*_.

### S3 Data and observational model

For inference, we, therefore, consider daily confirmed cases *c*_*i*_ of patients arriving at *H*^1^ and daily reported deaths *d*_*i*_, for the metropolitan area or region being analyzed.

The first default model for count data is a Poisson distribution; however, epidemiological data tends to be over disperse. Thus, an over disperse generalized Poisson distribution may be needed to correctly, and safely, model these types of data. Following [16] (see main paper) the NB distribution is re parametrized in terms of its mean *μ* and “overdispersion” parameters *θ* and *ω*_*NB*_, with 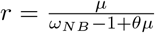 and 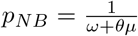, the number of failures before stopping and the success probability, respectively, in the usual NB parametrization. For data *y*_*i*_ we let *y*_*i*_ ∼ *NB*(*pμ*(*t*_*i*_), *ω, θ*), with fixed values for the overdispersion parameters *ω*_*NB*_, *θ* and an additional reporting probability *p*. The index of dispersion is *σ*^2^*/μ* = *ω*_*NB*_ + *θμ*. Over dispersion with respect to the Poisson distribution is achieved when *ω*_*NB*_ > 1 and the index of dispersion increases with size if *θ* ≠ 0; both desirable characteristics in outbreak data, adding variability as counts increase. In both cases we found good performance fixing *ω*_*NB*_ = 2. To model daily deaths, we fixed *θ* = 0.5 and for daily cases *θ* = 1 implying higher variability for the later. The reporting probabilities are 0.95 for deaths and 0.85 for cases, with the assumption that the *c*_*i*_’s are confirmed sufficiently severe cases arriving at hospitals. As explained in the main paper, the theoretical expectations estimated in terms of the dynamical model are given by *μ*_*D*_(*t*_*i*_) and *μ*_*c*_(*t*_*i*_) for dead and cases, respectively.

### S4 Modeling interventions and Bayesian Inference

We assume conditional independence in the data, and therefore from the NB model, we obtain a likelihood. Our parameters are the contact rate parameters *β*’s, the *ω*’s and crucially we also infer the initial conditions *E*(0), *I*^*A*^(0), *I*^*S*^(0). Letting *S*(0) = *N* − (*E*(0) + *I*^*A*^(0) + *I*^*S*^(0)) and setting the rest of the parameters to zero, we have all initial conditions defined and the model may be solved numerically to obtain *μ*_*D*_ and *μ*_*c*_ to evaluate our likelihood. We use the *lsoda* solver available in the *scipy*.*integrate*.*odeint* Python function.

Moreover, as explained in the main paper, we also estimate *ω*_*i*_ with *N*_*eff*_ = *ω*_*i*_*N*, both before and after a change point (“relaxation day”; we show intervention days with black vertical lines and relaxation change points with red vertical lines in our plots, see next section). To make it a bit simple, we also force the model to have a new *β* parameter after a relaxation day. However, the process of numerically solving the system of ODE’s is slightly more complex. For a set of parameter values, including the, to evaluate the likelihood, one needs to apply the solver from time *t* = 0 to the first relaxation day, considering *N*_*eff*_ = *ω*_1_*N*. Then for *ω*_2_ > *ω*_1_ we use the last values of all state variables as initial values for a second solve now with *N*_*eff*_ = *ω*_2_*N*, and so forth.

To sample from the posterior we resort to MCMC using the “t-walk” generic sampler [42]. The MCMC runs semi-automatic, with a fairly consistent burn-in of 1,000 iterations (sampling initial values from the prior). We perform subsampling using the Integrated Autocorrelation Time, with pseudo-independent sample sizes of 1,000 to 1,500 with 400,000 iterations of the MCMC. This process takes roughly 60 min in a 2.2 GHz processor.

To illustrate the whole posterior distribution, for any state variable *V* (or *μ*_*c*_(*t*_*i*_)), for each sampled initial conditions and *β*’s the model is solved at time *t*_1_, *t*_2_, …, *t*_*k*_, including possibly future dates, obtaining a sample of *V* (*t*_*i*_) values for each *t*_*i*_. The median and other desired quantiles are plotted vertically for each date considered, obtaining the plots as in Figure 3. Note that the traced median or other plotted quantiles do not necessarily correspond to any given model trajectory. It provides a far richer Uncertainty Quantification approach than the classical parameter estimates plug-in approach. Indeed, the sampled values for *V* (*t*_*i*_) do correspond to Monte Carlo samples of the posterior predictive distribution for *V* (*t*_*i*_).

### S5 Adding age structure

Adding group ages is straightforward in these types of models. The number *a* of age groups is decided, and our model is repeated *a* times. Different residence times may be included [50] but we preferred to concentrate on the different transition probabilities *g, h* that vary nearly two orders of magnitude using age groups [0, 25], (25, 50], (50, 65], (65, 100] [1]. The age structure is used to divide the initial infectious and susceptible population proportional to each age group size. We infer the same number of parameters, using a single *β*, with an optional weighting contact matrix [51] to model different contact rates among age groups in specific regions. Using a sufficiently flexible software design, progressing to an age-structure model is not complex; nonetheless, the MCMC may run substantially slower. We have experimented with our age-structured model using census data from Mexico and both uniform and non-uniform contact matrices. However, we do not report any of these results here, given that our non-age structure model has sufficiently enough predictive power, as already discussed.

### S6 Confounding effect of *N*_*eff*_ × *f*

To explain the confounding effect of *N*_*eff*_ × *f* we have two observations. First, if we let 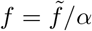 for some 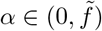 then differential equations for the variables *I*^*c*^, *H*^1^, *H*^2^, *H*^3^, *U* ^1^, *U* ^2^ and *D* remain invariant and the equations for *I*^*s*^ becomes

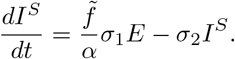

By letting 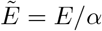 the equation for *I*^*s*^ is also invariant with the substitution of *E* by 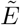. Now, the equation for 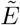 is given by

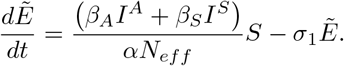

By letting *Ñ*_*ef f*_ = *αN*_*eff*_ the latter equations becomes also invariant under the substitution of *E* by 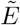. Therefore for the lower branch in the model (see Figure **??**) the system of equations is invariant under the change of *f* and *N*_*eff*_ by 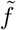 and *Ñ*_*ef f*_ provided 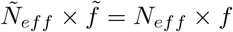 holds. We need to adapt the equations for *S, I*^*A*^, and *R* to get a consistent system of equations.

Second, to infer parameter *β* we inform the system with data at *H*^1^ and *D* compartments. If 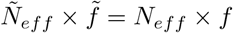 holds, in view of our first observation, to fit these data the fluxes *fσ*_1_*E* and 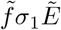 in either case have to be the same. The solutions in the compartment *I*^*S*^ and after do not change in this case, but the individuals in the *I*^*A*^ compartment does change depending on which combination of *N*_*eff*_ and *f* or *Ñ*_*ef f*_ and 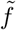 is considered. There is a range of validity for *α* where the inference of *β* does not change, but we do not explore this property further.

**Fig S6.**
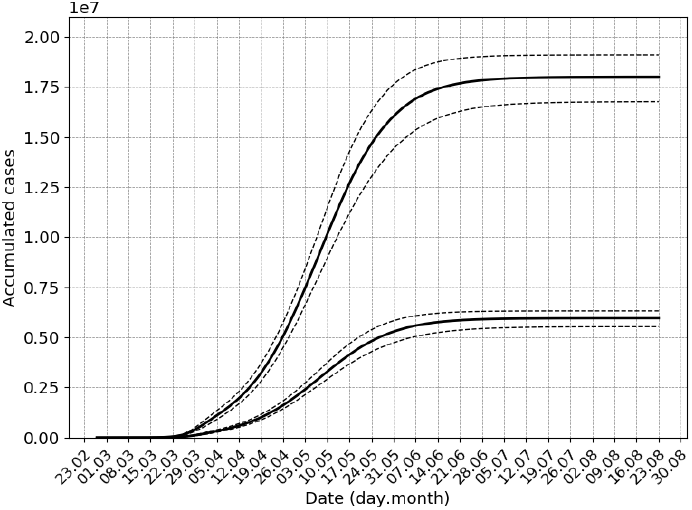
Outbreak analysis for Mexico city metropolitan area, data until 15 May. Total number of recovered *R*; with *N*_*eff*_ = *N*, total population, and *f* = 0.05, *R*(∞) reaches approximately 17.5 10^6^ while with *N*_*eff*_ = *N/*3, *f* = 0.05 · 3, *R*(∞) only reaches roughly 5.5 10^6^. However, the fit for cases and deaths and the predictive curves for hospital demand are identical (results not shown).

We also present numerical simulations to confirm this confounding effect (see Figure S6), but until the asymptomatic infection is fully described, it is not possible to resolve this issue.

### S7 Data sources

We take metropolitan areas delimitation and population from [52] and [53], respectively. Official records of COVID-19 confirmed cases and deaths are reported in [54].

